# The influence of comorbidity on the severity of COVID-19 disease: A systematic review and analysis

**DOI:** 10.1101/2020.06.18.20134478

**Authors:** Nazar Zaki, Elfadil A. Mohamed, Sahar Ibrahim, Gulfaraz Khan

## Abstract

**Background:** A novel form of coronavirus disease (SARS-CoV-2) has spread rapidly across the world. What risk factors influence the severity of the disease is of considerable importance.

**Aim:** This research offers a systematic review and meta-analysis of the correlation between common clinical conditions and comorbidities and the severity of COVID-19.

**Methodology:** Two independent researchers searched Europe PMC, Google Scholar, and PubMed databases for articles related to influence comorbidities have on the progress of the disease. A search engine was also created to screen a further 59,000 articles in COVID-19 Open Research Dataset (CORD-19). Random-effects modeling was used to pool 95% confidence intervals (CIs) and odds ratios (ORs). The significance of all comorbidities and clinical conditions to the severity of the disease was evaluated by employing machine-learning techniques. Publication bias was assessed by using funnel-plots and Egger's test. Heterogeneity was tested using I^2^.

**Results:** The meta-analysis incorporated 12 studies spanning 4,101 confirmed COVID-19 patients who were admitted to Chinese hospitals. The prevalence of the most commonly associated co-morbidities and their corresponding odds ratio for disease severity were as follows: coronary heart disease (OR 2.97 [CI: 1.99-4.45], p < 0.0001), cancer (OR 2.65 [CI: 1.12-6.29], p < 0.03), cardiovascular disease (OR 2.89 [CI: 1.90-4.40], p < 0.0001), COPD (OR 3.24 [CI: 1.66-6.32], p = 0.0), and kidney disease (OR 2.2.4 [CI: 1.01-4.99], p = 0.05) with low or moderate level of heterogeneity. The most frequently exhibited clinical symptoms were fever (OR 1.37 [CI: 1.01-1.86], p = 0.04), myalgia/fatigue (OR 1.31 [CI: 1.11-1.55], p = 0.0018), and dyspnea (OR 3.61, [CI: 2.57-5.06], p = <0.0001). No significant associations between disease severity and liver disease, smoking habits, and other clinical conditions, such as a cough, respiratory/ARDS, diarrhea or chest tightness/pain were found. The meta-analysis also revealed that the incubation period was positively associated with disease severity.

**Conclusion:** Existing comorbidities, including COPD, cardiovascular disease, and coronary heart disease, increase the severity of COVID-19. Some studies found a statistically significant association between comorbidities such as diabetes and hypertension and disease severity. However, these studies may be biased due to substantial heterogeneity.

## 1. Introduction

The 2019/2020 emergence of the novel COVID-19 disease and its swift global expansion represents a health emergency for all of humanity. This novel virus, which causes severe acute respiratory disease, is believed to be a member of the same family of coronaviruses as the Severe Acute Respiratory Syndrome coronavirus (SARS-CoV) and the Middle East Respiratory Syndrome coronavirus (MERS-CoV) [1]. Unprecedented efforts are currently being made to identify the risk factors that increase the severity of the disease. The World Health Organisation (WHO) has suggested that patients with certain medical conditions and elderly people are at greatest risk of developing more severe disease [2]. According to reports from Johns Hopkins University and as of July 30, 2020, over 17 million people have been infected and over 667,000 have died from the disease. The majority of deaths are thought to have associations with the existence of one or more comorbidity [3]. In general, it is believed that patients with compromised immune system are especially at high risk [4]. A number of researchers have examined the clinical/epidemiological characteristics of patients suffering COVID-19, but there has been insufficient investigation regarding mortality risk factors [5]. The identification of the principal risk factors and employing appropriate clinical interventions to mitigate them could save numerous lives. Numerous studies indicate that kidney disease, Chronic Obstructive Pulmonary Disease (COPD), coronary heart disease, liver disease, cardiovascular disease, diabetes mellitus, and hypertension are amongst the most important risk factors for COVID-19. This paper will undertake a systematic of the literature to illuminate the way in which the impact of the COVID-19 virus can be made more severe by the existence of such comorbidities and extant conditions.

## 2. Materials/methodology

### 2.1. Protocol and registration

The protocol in this paper was drafted using the Preferred Reporting Items for Systematic Reviews and Meta-analysis Extension for Scoping Reviews (PRISMA-ScR) [6]. The scoping review is a type of knowledge synthesis, that follows, a systematic approach to map pieces of evidence, identify the main concepts that underpin a topic; and determine where the gaps are.

### 2.2. Eligibility criteria

In this research, we looked for all relevant published articles between the dates of December 1, 2019, and May 20, 2020, i.e. those that were related to comorbidity and its influence on the severity of COVID- 19. Papers were excluded if they were related to the pediatric patients, if they were not written in English, if they had not been peer reviewed, or if they were not original (e.g. reviews, editorials, letters, commentaries, or duplications). Papers mainly focused on severe vs non-severe groups of patients were considered. Severe group are those who develop severe illnesses. These cases are more likely to result in ICU admission or death.

### 2.3. Information sources

An automated search was undertaken on the COVID-19 Open Research Dataset (CORD-19), which is regularly updated [7]. At the time, this dataset contained more than 128,000 scholarly articles; 59,000 of which included full text that related to coronavirus, SARS-CoV-2, and/or COVID-19. Moreover, an independent search was undertaken to systematically examine relevant articles in other databases such as PubMed, Europe PMC, and Google Scholar.

### 2.4. Search

A search engine running over the BM25 search index was created so that all articles in this dataset could be screened [8]. The BM25 retrieval function creates a document ranking for a dataset on the basis of whether or not searched terms appear in the documents, no matter how proximate they may be to each other. Indexing of the papers occurs through simple application of preprocessing functions that clean and tokenize abstracts. Having indexed all documents, document vectors were created through loading optimized cached JSON tokens and subsequent application of a document similarity index founded on Annoy [9]. Annoy is a C++ library that has Python bindings for searching for documents within a space that make a close match to a specific query. This is an efficient and simple process as it creates substantial read-only file-based data structures used with memory mapping allowing several processes to work with identical data. More detail on this implantation can be found at HTTP://github.com/dgunning/cord19.

Articles were then filtered employing questions and keywords such as “Effects of diabetes on COVID/coronavirus/SARS-CoV-2/nCoV/COVID-19 disease severity?” This search was undertaken for every comorbidity and symptom under consideration in this research (Fig. 1).

**Fig. 1:**
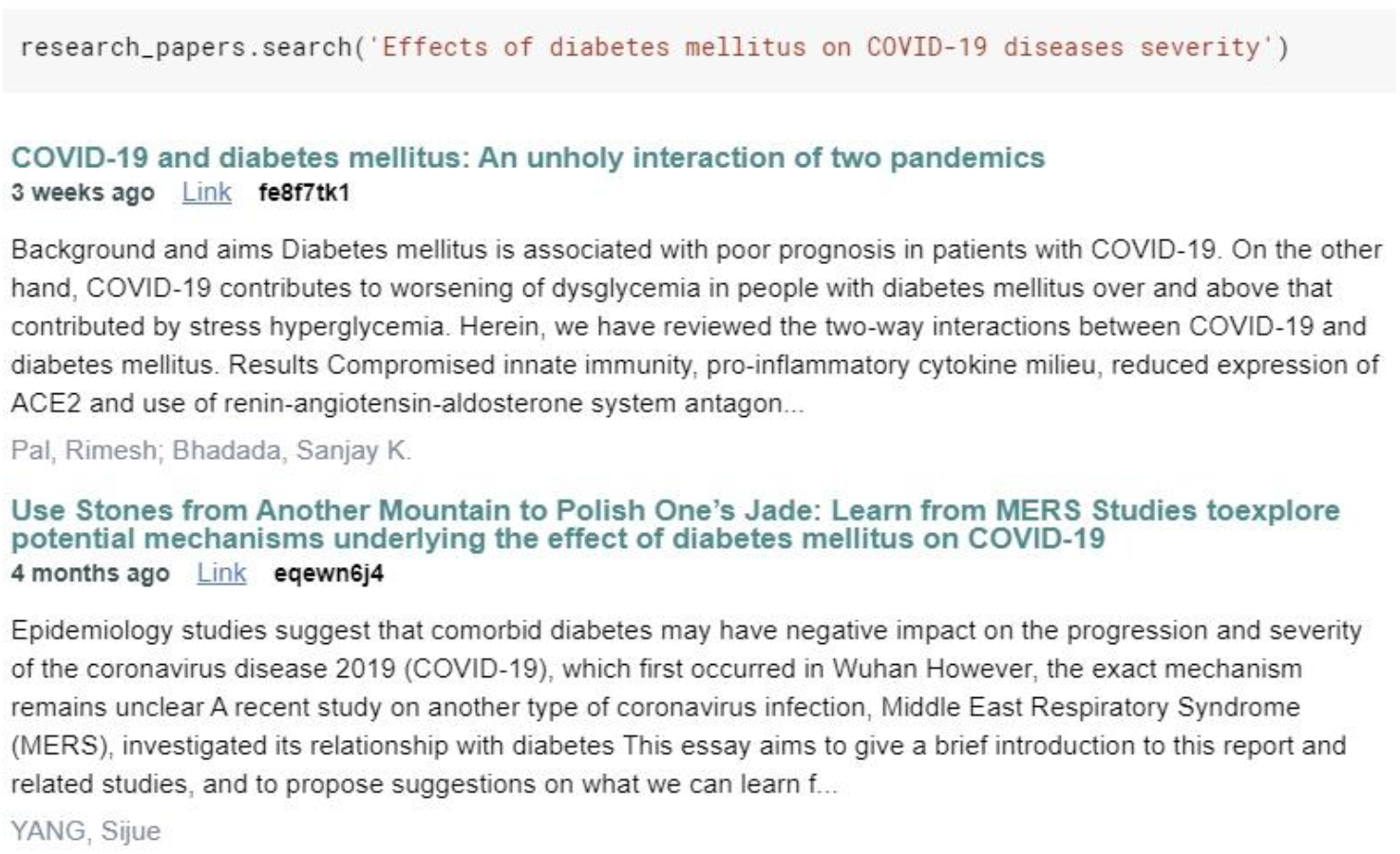
Example of retrieved articles from a search for diabetes mellitus

### 2.5. Selection of evidence sources

In addition to the automated search conducted using the search engine, an independent search was undertaken by two reviewers (NZ and EA), who examined the PubMed, Europe PMC, and Google Scholar databases. To increase consistency, the reviewers screened the automated and manually extracted publications and discussed the results before beginning the screening for the review. Both reviewers sequentially evaluated the titles, abstracts, and full text of all publications identified during the automated and manual searches to identify potentially relevant publications. The articles that remained following the screening process underwent independent screening a second time by the two reviewers (NZ and EA). Any disagreement was settled either by consensus or by the casting vote of a third reviewer (GK). The research was completed on May 20^th^, 2020.

### 2.6. Data charting process

As a means of identifying the variables of interest for the purpose of the study, a data-charting form was jointly developed by the two reviewers (NZ and EA). The two reviewers independently charted the data, discussed the results, and continually updated the data-charting form. The charting table was further refined during the review stage and updated accordingly.

### 2.7. Data items

Data was compiled on paper reference number, author name, publication venue, month and year of publication, basic information (number of patients, median age, number of male and female under severe/non-severe conditions), number of severe/non-severe COVID-19 patients with comorbidities (diabetes, hypertension, coronary heart disease, cancer, liver disease, COPD, kidney disease, cardiovascular disease), number of severe/non-severe COVID-19 patients with symptoms (cough, respiratory/ARDS, fever, myalgia/fatigue, dyspnea, diarrhea, chest tightness/pain), and other information such as incubation period, smoking status, and deaths). Where either vital or non-vital information was absent, requirement for admission to ICU was considered indicative of the severity of the patient's condition.

### 2.8. Critical appraisal of individual sources of evidence

Meta-analysis was undertaken by employing a Python library “meta”. The odds ratios for developing severe disease was estimated for each of the identified co-morbidities for patients with severe disease compared to those with non-severe form of the disease. Due to internal and external heterogeneities, random effect modeling was employed to estimate the average effects and their precision, offering a more cautious estimate regarding the 95% confidence intervals (CIs). Statistical heterogeneity was assessed using the I^2^ statistic. The study was performed according to the process outlined in the Cochrane Handbook for Systematic Reviews of Interventions Version 6.0 [10]. Specially, a score of 0% to 40% was considered to be unimportant, 30% to 60% was considered to represent moderate heterogeneity, 50% to 90% was considered to represent substantial heterogeneity, and 75% to 100% was considered considerable heterogeneity.

Machine learning techniques based on regression, e.g., support vector machine (SVM) [11], linear regression, multi-perceptron [12], random forest [13], and attribute selection techniques, such as Classifier Subset Evaluator, Correlation Ranking Filter, and Relief Ranking Filter [14] as implemented in WEKA [15], were employed to determine how useful and significant different variables were in terms of the prediction of levels of severe instances of COVID-19. Consideration was only given to kidney disease, coronary heart disease, COPD, liver disease, cardiovascular disease, cancer, diabetes mellitus, and hypertension conditions. Clinical symptoms that were added for consideration as independent attributes were diarrhea, chest tightness/pain, respiratory failure, dyspnea, fatigue, coughing, fever, and incubation period (the time from the probable earliest contact with a source of transmission and the earliest recognition of the first symptoms [1]), current smoking status, and patient gender.

### 2.9. Synthesis of the results

The studies were grouped by comorbidity: diabetes, hypertension, coronary heart disease, cancer, liver disease, COPD, kidney disease, and cardiovascular disease. The clinical conditions of interest were cough, respiratory/ARDS, fever, myalgia/fatigue, dyspnea, diarrhea, and chest tightness/pain. In addition, incubation period, and smoking habits were also included in the data set.

## 3. Results

### 3.1. Selection of evidence sources

As shown in Fig. 2, 303 articles were identified as being relevant to the objectives of this study as a result of the search of Europe PMC (n = 110), PubMed (n = 44), Google Scholar (n = 81), and the COVID- 19 database (n = 68); 108 of these were removed as they were duplicates. In total, 195 text articles were screened against the eligibility criteria. A further 125 articles were removed on the basis that they were not English-language articles, had not been peer-reviewed, did not contain full text, were irrelevant, were duplicates, or took the form of editorial notes.

After the full-text article assessment, an additional 58 articles were removed because they were irrelevant (n = 6), severity of the disease was not mentioned (n = 5), incomplete data were available for calculating OR and other statistics (n = 16), the patient population was not suitable (n = 4), the study design was not suitable (e.g., editorial, narrative review, guideline, commentary, letter, study without primary clinical data) (n = 24), or they were duplicates of previously included articles (n = 2). Ultimately, the meta-analysis group consisted of 12 articles that met all quantitative and qualitative standards.

**Fig. 1.**
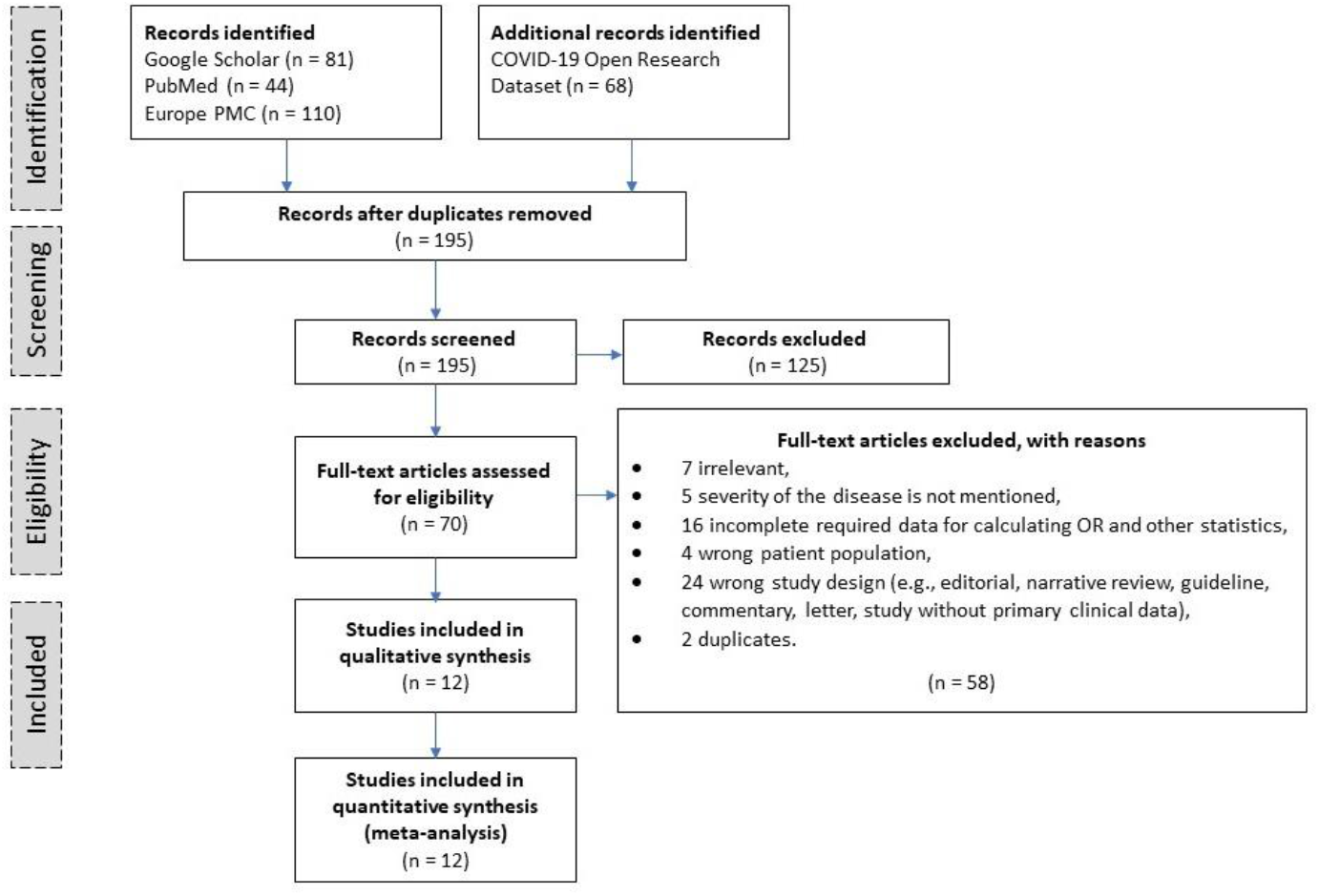
Flowchart of screening process outcomes during the different phases of the systematic review.

### 3.2. Results of individual evidence sources

As can be observed in Table 4, the data of interest spanned 4,101 patients, of which 1,179 had a severe form of COVID-19 and 2,922 had a non-severe form of COVID-19. The median age of the population was 58.8 years for the severe cases and 48.5 years for the non-severe cases. 53.6% of patients were male. The meta-analysis demonstrated that the highest levels of comorbidity were associated with hypertension (22.1%), diabetes (11.3%), cardiovascular disease (10.8%), liver disease (6.3%), coronary heart disease (5.5%), cancer (4.4%), kidney disease (3.8%), and COPD (2.5%). The most common clinical symptoms were fever (74.5%), cough (62.2%), myalgia/fatigue (38.8%), dyspnea (33.9, respiratory failure/ARDS (20.6%), diarrhea (11.2%), and chest tightness/pain (16.8%). It was also noted that comorbidity, such as coronary heart disease, cancer, COPD, kidney disease, and cardiovascular disease, was associated with disease severity, as indicated by the p-value of ≤ 0.05 (statistically significant) and the low or moderate heterogeneity. Similarly, symptoms such as fever, myalgia/fatigue, and dyspnea were also positively correlated with disease severity, as too was incubation period.

### 3.3. Synthesis of the results

Figure 3 (A-H) presents an analysis of the comorbidities across the severe and non-severe COVID-19 groups. Although the meta-analysis indicated that disease severity was associated with diabetes (OR 2.27, [95% CI: 1.46-3.53], p = 0.0; I^2^: 62%), and hypertension (OR 2.43 [95% CI: 1.71-3.45], p < 0.0001); I^2^:62%), these findings should be treated with caution because the level of heterogeneity in both cases was substantial (62%). On the other hand, the meta-analysis indicated that disease severity was statistically significantly associated with coronary heart disease (OR 2.97 [95% CI: 1.99-4.45], p < 0.0001; I^2^: 0%), cancer (OR 2.65 [95% CI: 1.12-6.29], p < 0.03; I^2^: 0%), cardiovascular disease (OR 2.89 [95% CI: 1.90-4.40], p < 0.0001, I^2^:38%), COPD (OR 3.24 [95% CI: 1.66-6.32], p = 0.0; I^2^: 0%), and kidney disease (OR 2.2.4 [95% CI: 1.01-4.99], p = 0.05, I^2^: 38%) with a low or moderate level of heterogeneity. The I^2^ test for heterogeneity ranged between 0% to 38%, indicating a fair level of statistical heterogeneity. For coronary heart disease, cancer, COPD and coronary heart disease, I^2^ was equal to 0%, meaning no heterogeneity was identified. This indicates that this study was adequately homogenous. However, the results related to cancer and kidney disease should be treated with caution because the forest-plots revealed that the overall effect estimate was touching the “no effect” line. Moreover, no significant correlation was found between disease severity and liver disease.

Figure 4 (A-G) indicates that the associations between symptoms and COVID-19 severity were as follows: with fever (OR 1.37, 95% CI: 1.01-1.86, p = 0.04; I^2^: 40%), myalgia/fatigue (OR 1.31, 95% CI: 1.11-1.55, p = 0.0; I^2^: 0%), and dyspnea (OR 3.61, 95% CI: 2.57-5.06, p = <0.0001; I^2^: 43%). However, the fever results should be treated with caution because the forest-plots revealed that the overall effect estimate was touching the “no effect” line.

Although the associations between COVID-19 severity and respiratory failure/ARDS (OR 11.46, 95% CI: 3.24-40.56, p = 0.0; I^2^: 89%), and chest tightness/pain (OR 2.17, 95% CI: 1.40-3.36, p = 0.0; I^2^: 71%) were statistically significant, the meta-analysis indicated substantial heterogeneity.

**Fig. 3.**
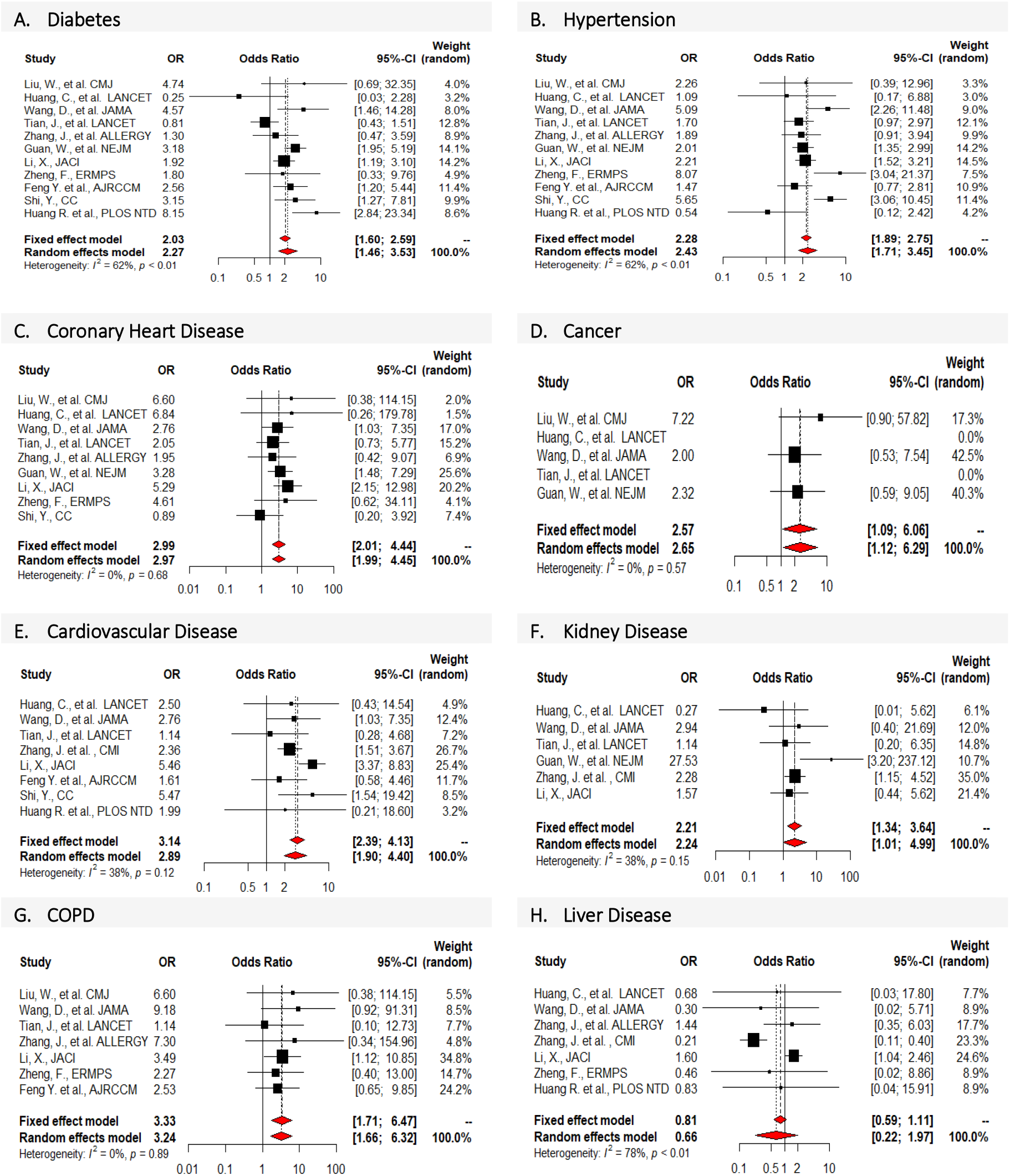
Forest-plots indicate the comorbidities under investigation (coronary heart disease, cancer, cardiovascular disease, kidney disease, and COPD) and different levels of associations with poor composite outcomes, severe COVID-19, and ICU care requirements. This did not hold true for liver disease patients.

**Fig. 4.**
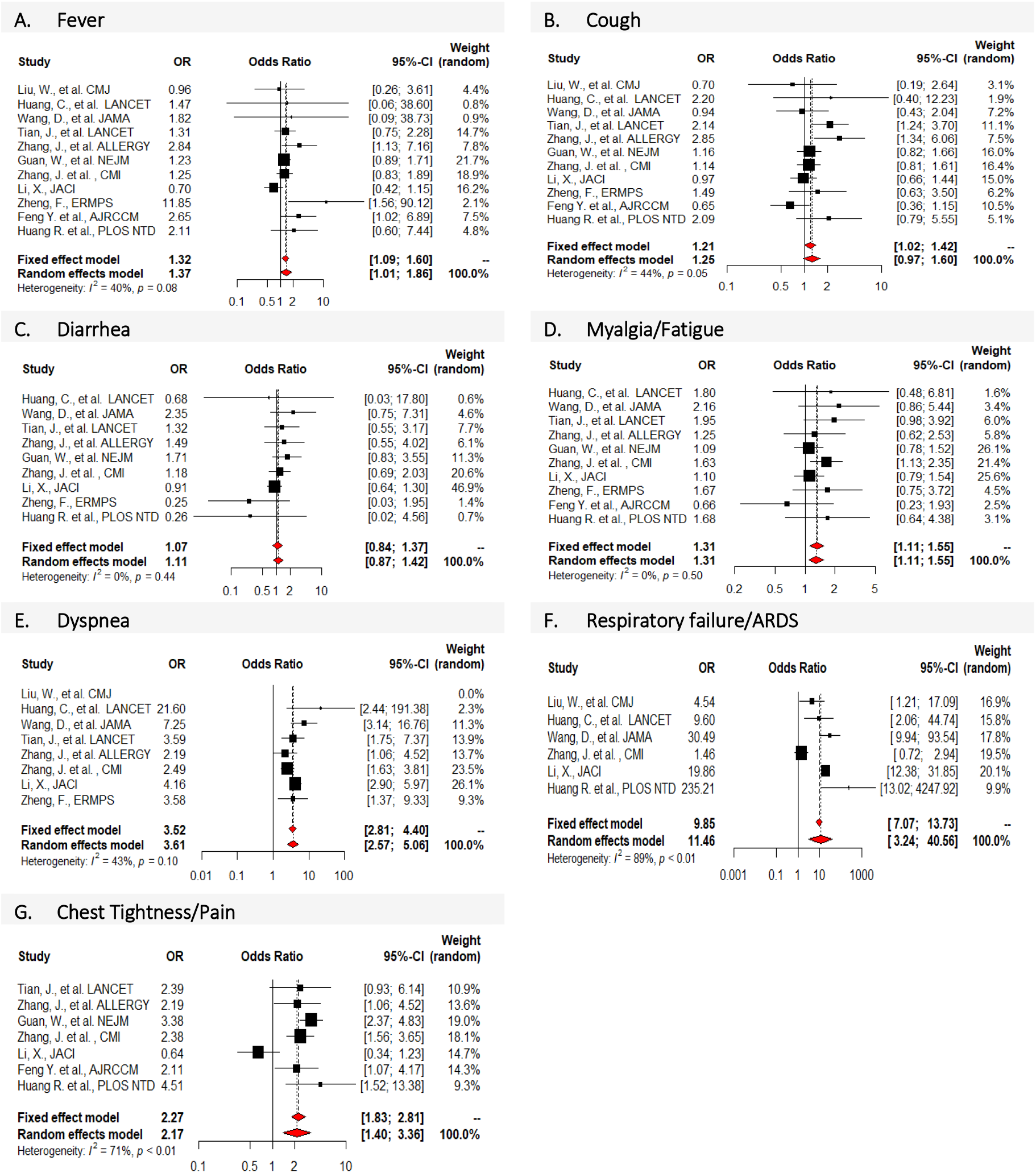
Forest-plots indicate that clinical conditions like myalgia/fatigue, fever, and dyspnea, had associations with higher composite poor outcomes, severe COVID-19, and ICU care requirements. This did not hold true for patients suffering from symptoms such as cough, respiratory/ARDS, diarrhea, and chest tightness/pain.

To evaluate publication bias as part of the meta-analysis, we employed the R meta-package to visualize the funnel plots. Funnel plots are often used as an exploratory tool to visually assess the possibility of publication bias within the meta-analysis. However, in some cases, the funnel plot may not be highly informative, and it may be challenging to spot asymmetries when less than 10 studies are considered. Therefore, an Egger's regression test was also included to provide more evidence to facilitate an evaluation of publication bias. The outcomes of the publication bias assessments are shown in Figs. 5 and 6. Visually, the figures show only minor asymmetry (with or without inclusion of the studies lacking individual participant data), and the Egger's test for asymmetry was not significant (p-value > 0.05). Thus, a publication bias mechanism was not a major cause for concern in this study. However, in the case of cancer association, for instance, only three studies qualified for the analysis; as such, the publication bias evaluation may not be meaningful. Two studies were limited to cancer patients. These were automatically eliminated from the meta-analysis.

**Fig. 5:**
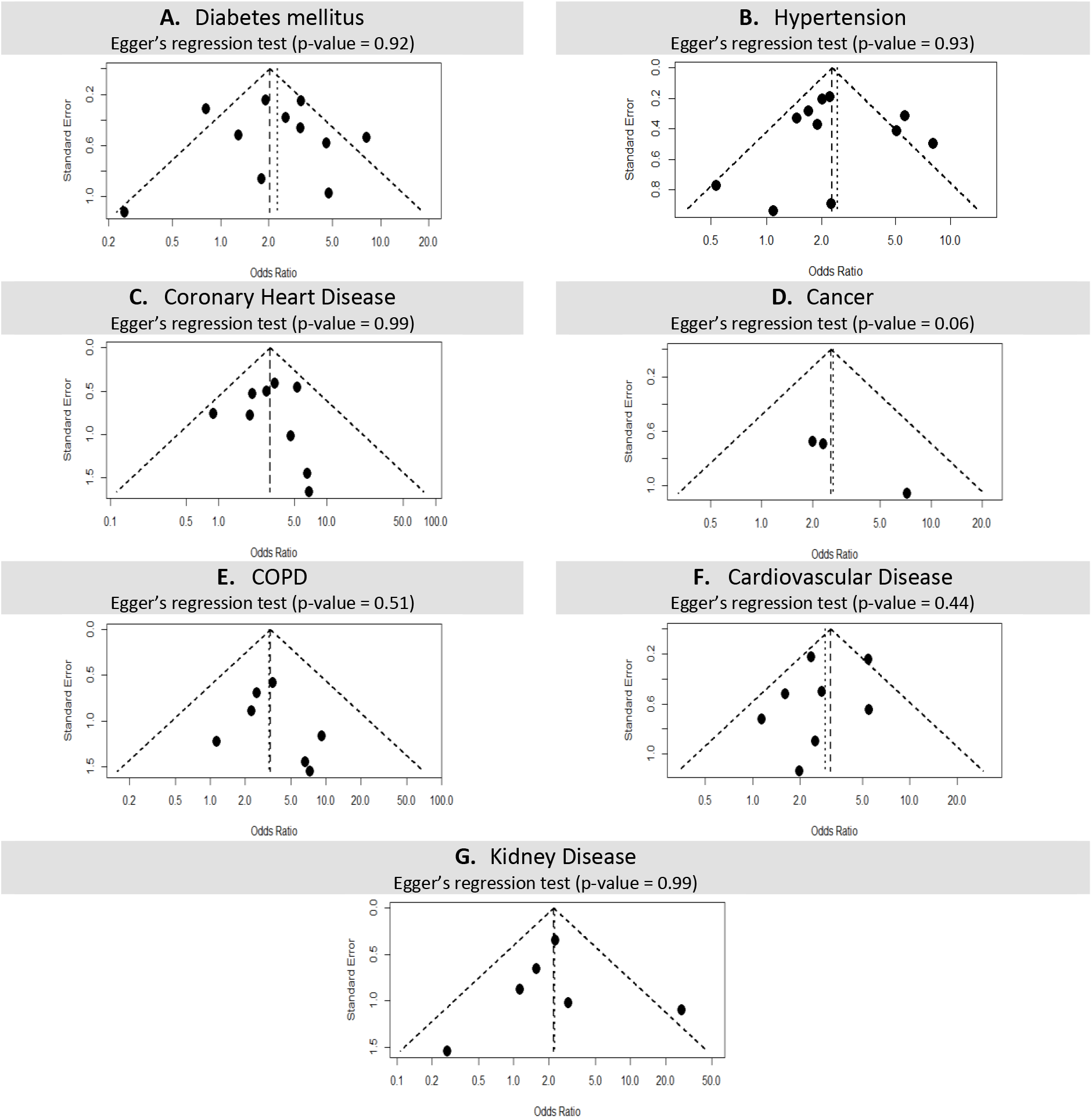
Publication bias funnel plots for comorbidities with significant associations (p-value < 0.05), which included diabetes mellitus, hypertension, coronary heart disease, cancer, COPD, cardiovascular disease, and kidney disease. Visually, the figures showed only minor asymmetry, and the Egger's regression test for asymmetry was not significant (p-value > 0.05). Thus, the publication bias mechanism was not a major cause for concern in this study.

**Fig. 6:**
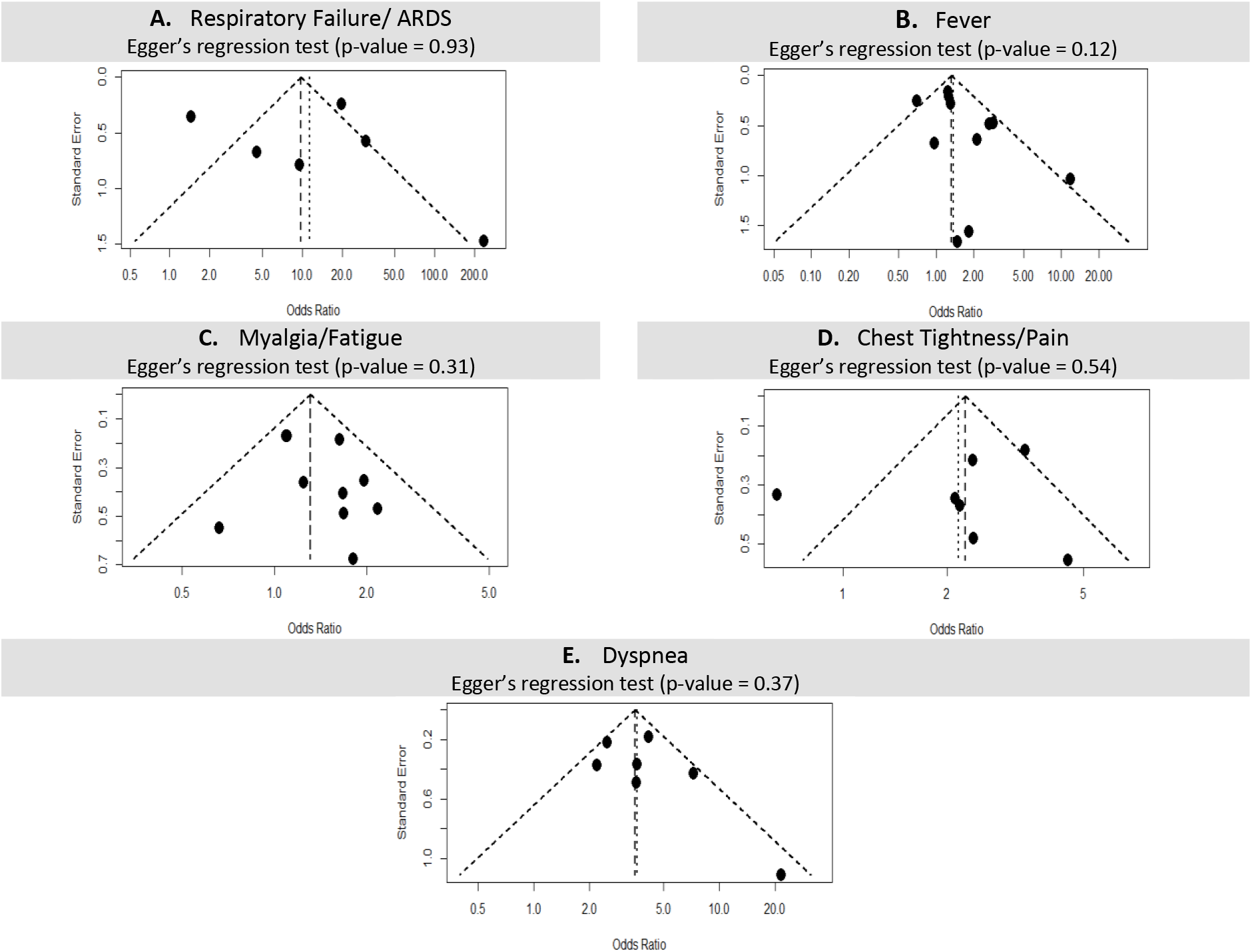
Publication bias funnel plots for conditions with significant associations (p-value < 0.05), including respiratory failure/ ARDS, fever, myalgia/fatigue, chest tightness/pain and dyspnea. Visually, the figures showed only minor asymmetry, and the Egger’s regression test for asymmetry was not significant (p-value > 0.05). Thus, a publication bias mechanism was not a major

This analysis employed the seven-machine learning/feature selection techniques outlined in the methodology section. The results indicated that coronary heart disease was listed in the top 10 crucial attributes for all seven techniques, diabetes (6/7), hypertension (4/7), and COPD (4/7). Clinical conditions like fever (5/7), chest pain (4/7), and fatigue (4/7) were similarly listed in the top 10 important attributes. Fig. 7 presents the intersections of comorbidities and clinical conditions listed in the top 15 crucial attributes yielded by four feature selection techniques; namely, SMO (Support Vector Machine), Relief (Relief Ranking Filter), Corr-attr (Correlation Ranking Filter), and ClassifierAttr (Classifier Subset Evaluator). All four listings included comorbidities such as coronary heart disease, diabetes, COPD, hypertension, and cardiovascular disease besides myalgia/fatigue as a more likely clinical condition.

**Fig. 7:**
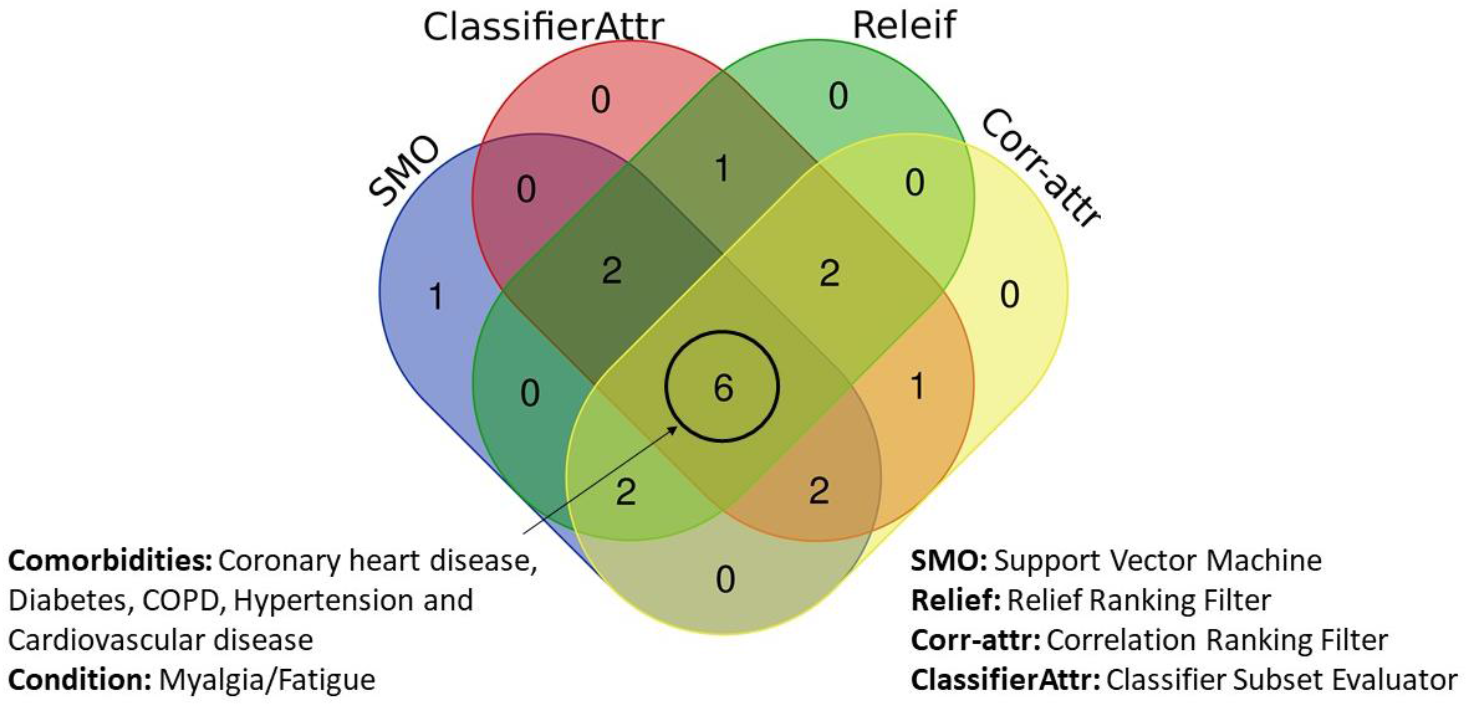
Six comorbidities (coronary heart disease, diabetes, COPD, hypertension, and cardiovascular disease) and one clinical condition (myalgia/fatigue) were in the top selected attributes associated with COVID-19 severity.

## 4. Discussion

### 4.1. Summary of evidence

This study involved a comprehensive vision review/meta-analysis of the correlation between comorbidities, e.g., diabetes and hypertension, and the severity of COVID-19. The meta-analysis was based on data from 12 studies spanning 4,101 patients who were admitted to Chinese hospitals with a COVID-19 diagnosis confirmed by laboratory tests. Patients with a severe form of the disease had a median age of 58.8, while those with a non-severe form of the disease had a median age of 48.5. There were more males than females in the study (2,196 males, 1,702 females). Two of the studies ([23] and [28]) found a significant correlation between gender and disease severity. Li et al. [23] concluded that severe male patients with heart injury, hyperglycemia, and high-dose corticosteroid use may have a high risk of death. Channappanavar et al. [28] conducted experimental works on mice and found that male mice were more susceptible to SARS-CoV infection than age-matched females. They concluded that males have a higher likelihood of infection due to higher exposure levels.

The meta-analysis of comorbidity found that diabetes mellitus (11.3%) and hypertension (22.1%) were the two most prevalent conditions in the patient cohort; this almost exactly parallels the levels of hypertension and diabetes in the population of China as a whole (hypertension 23.2% [29], diabetes mellitus 10.9% [30]). However, we must treat these results with caution because the level of heterogeneity in both cases was substantial (62%).

The meta-analysis also demonstrated that unfavorable outcomes had significant associations with diabetes (OR 2.27, [95% CI: 1.46-3.53], p = 0.0), hypertension (OR 2.43 [95% CI: 1.71-3.45], p < 0.0001)), coronary heart disease (OR 2.97 [95% CI: 1.99-4.45], p < 0.0001), cardiovascular disease (OR 2.89 [95% CI: 1.90-4.40], p < 0.0001, I^2^:38%), cancer (OR 2.65 [95% CI: 1.12-6.29], p < 0.03; I^2^: 0%), COPD (OR 3.24 [95% CI: 1.66-6.32], p = 0.0; I^2^: 0%), and kidney disease (OR 2.2.4 [95% CI: 1.01-4.99], p = 0.05, I^2^: 38%) with a low or moderate level of heterogeneity. Fang et al. [31] found that human pathogenic coronaviruses, e.g., SARS-CoV and SARS-CoV-2, achieve binding with target cells through the use of Angiotensin-Converting Enzyme 2 (ACE2), which originates from epithelial cells in the blood vessels, kidneys, intestines, and lungs [32]. ACE2 levels are considerably higher for patients suffering from Type I or Type II diabetes. As such, they are treated with ACE inhibitors and/or Angiotensin II Type-1 Receptor Blockers (ARBs). These medications are also used to treat hypertension, and this causes an upregulation of ACE2 [33]. ACE2 may also increase following the use of ibuprofen or thiazolidinediones. Higher levels of ACE2 can, therefore, promote COVID-19 infection. Thus, we see that the evidence supports a mechanism for treating hypertension or diabetes with ACE2-stimulators could predispose patients to more severe COVID-19. Therefore, we concur with Fang et al. [31] that patients with diabetes, hypertension, or cardiac disease who are prescribed ACE2-increasing medication must be regarded as having a greater risk of developing severe COVID-19 and, as such, they should be closely monitored for the effects of self-medication.

In a recent review, Zhao et al. [34] found that extant COPD makes it four times more likely that severe COVID-19 will develop. They found no significant correlation between smoking and COVID-19 severity. However, smoking is harmful to the immune system and can undermine the body’s ability to respond to infection; as such, smokers are more susceptible to contracting infections [35]. This accords with the findings of this meta-analysis presented in Table 1 which indicates that there was no significant correlation (p-value = 0.1) between smoking habits and the severity of the COVID-19 disease in the data examined as part of the current study.

**Table 1:**
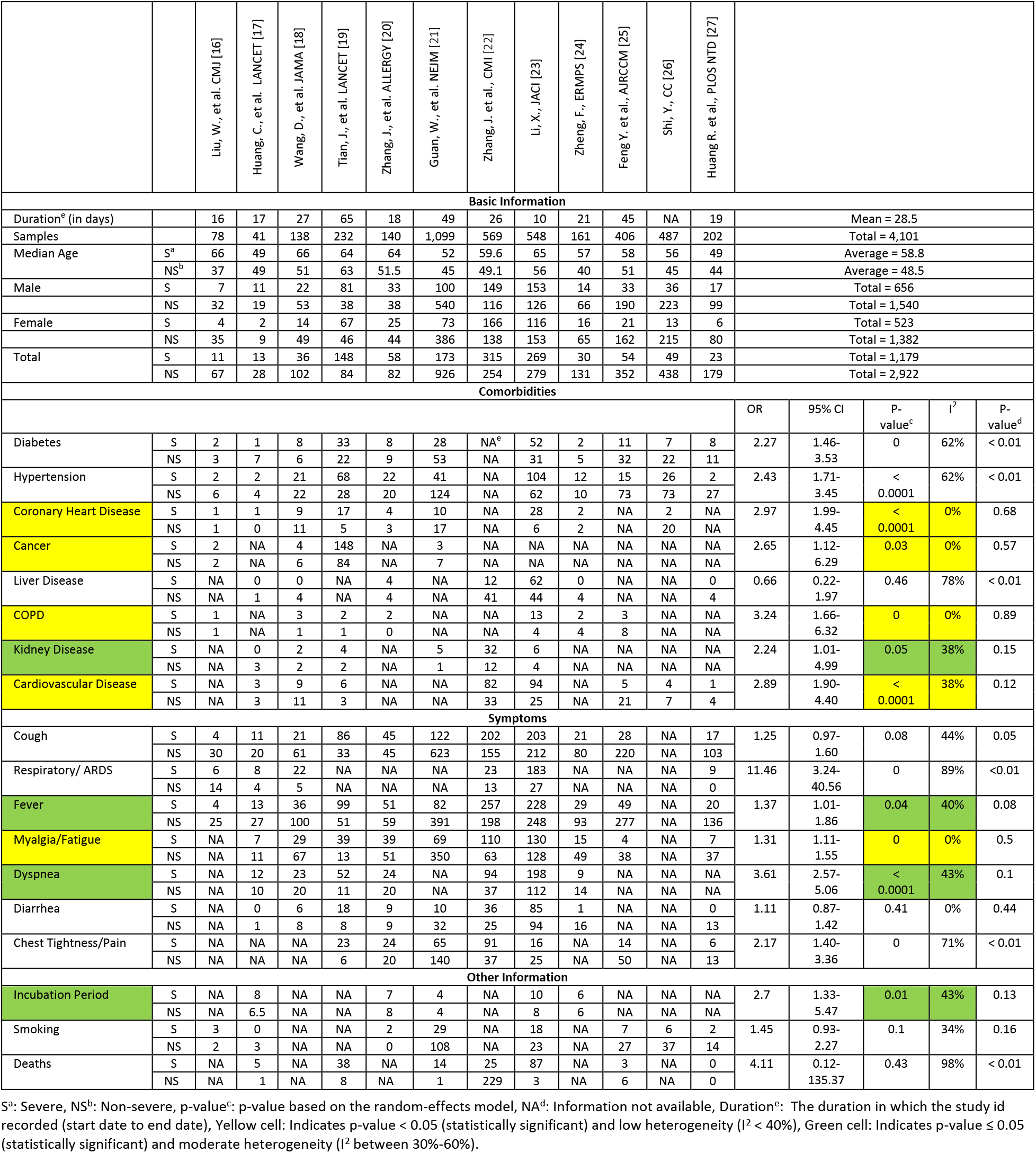
Summary of research used for meta-analysis including characteristics of severe/non-severe groups and number of common comorbidities and clinical conditions.

While patients exhibiting abnormal liver results are regarded as being significantly more likely to develop severe pneumonia, and employing ritonavir or lopinavir makes patients four times more likely to suffer liver injury [36], this meta-analysis did not find a significant correlation between liver disease and negative outcomes. This finding is aligned with recent research that was performed by [37], which found that, while it is common for COVID-19 patients to exhibit abnormal liver function index, it is not a significant factor in COVID-19 patients and may not be associated with serious negative clinical outcomes.

The study by Guan et al. [21] was the only one of the 12 articles under review that found a significant correlation between kidney disease and COVID-19 severity. When the articles are examined in combination, a minimum correlation was found (p = 0.05).

The clinical symptoms that were most frequently described in the papers included in this analysis were fever (74.5%), cough (62.2%), myalgia/fatigue (38.7%), dyspnea (33.9%), respiratory failure/ARDS (20.6%), diarrhea (11.2%), and chest tightness/pain (16.8%). The meta-analysis indicated that there was a significant association between COVID-19 severity and clinical conditions such as fever (OR 1.37, 95% CI: 1.01-1.86, p = 0.04), myalgia/fatigue (OR 1.31, 95% CI: 1.11-1.55, p = 0.0), and dyspnea (OR 3.61, 95% CI: 2.57-5.06, p = <0.0001). These findings are in agreement with the majority of published research. In terms of heterogeneity, in the majority of cases, zero or low heterogeneity was displayed by the I. As such, with the exception of diabetes, hypertension, and liver disease, conditions such as cough, respiratory/ARDS, diarrhea, and chest tightness/pain, exhibited relative homogeneity in all cases examined for this research.

### 4.2. Limitations

This scoping review and meta-analysis have certain limitations that should be addressed within future studies. First, only patients admitted to Chinese hospitals were included in the studies under review. As such, the findings cannot be generalized across a wider population. Second, the presence of multiple comorbidities within single patients was not considered (a limitation that is consistent across the existing studies in this context). Third, the results are only up to date as of May 20^th^, 2020. Due to the rapidly changing nature of the COVID-19 pandemic, significant additional data has been made available since that date. Finally, this meta-analysis does not include laboratory, radiographic, clinical, or demographic data.

### 4.3. Conclusions

Existing comorbidity, including coronary heart disease, COPD, and cardiovascular disease, along with clinical conditions, such as myalgia/fatigue and dyspnea, are associated with a higher risk of patients developing a more severe form of COVID-19. As more evidence becomes available in the shape of trustworthy published results from other regions/nations, further studies should be undertaken in this area.

## Data Availability

This is a literature review paper.

## Funding

Self-funding.

## Declarations

### Ethics approval and consent to participate

Ethical approval is not needed.

### Consent for publication

Not applicable.

### Availability of data and materials

Not applicable.

### Competing interests

The authors declare no competing or conflict of interest.

### Authors’ contributions

NZ and EA performed the literature scan and review, NZ performed the meta-analysis, all authors contributed to the paper writing, GK and SI validated the findings.

## Acknowledgements

The authors would like to thank Mr. Dwight Gunning for his kind assistance with the search engine. The authors would like to acknowledge the encouragement and support provided by Ajman University and United Arab Emirates University for conducting this research work.

## Notes

### Competing Interest Statement

The authors have declared no competing interest.

### Funding Statement

Self funded.

### Author Declarations

This is a literature review paper.

### Summary of Updates

Major work is done to improve the paper including the protocol in this paper which now drafted using the Preferred Reporting Items for Systematic Reviews and Meta-analysis Extension for Scoping Reviews (PRISMA-ScR). The results, figures, table and analysis are also revised and improved.

